# Not the Models You Are Looking For: Traditional ML Outperforms LLMs in Clinical Prediction Tasks

**DOI:** 10.1101/2024.12.03.24318400

**Authors:** Katherine E. Brown, Chao Yan, Zhuohang Li, Xinmeng Zhang, Benjamin X. Collins, You Chen, Ellen Wright Clayton, Murat Kantarcioglu, Yevgeniy Vorobeychik, Bradley A. Malin

## Abstract

**Objectives:** To determine the extent to which current Large Language Models (LLMs) can serve as substitutes for traditional machine learning (ML) as clinical predictors using data from electronic health records (EHRs), we investigated various factors that can impact their adoption, including overall performance, calibration, fairness, and resilience to privacy protections that reduce data fidelity.

**Materials and Methods:** We evaluated GPT-3.5, GPT-4, and ML (as gradient-boosting trees) on clinical prediction tasks in EHR data from Vanderbilt University Medical Center and MIMIC IV. We measured predictive performance with AUROC and model calibration using Brier Score. To evaluate the impact of data privacy protections, we assessed AUROC when demographic variables are generalized. We evaluated algorithmic fairness using equalized odds and statistical parity across race, sex, and age of patients. We also considered the impact of using in-context learning by incorporating labeled examples within the prompt.

**Results:** Traditional ML (AUROC: 0.847, 0.894 (VUMC, MIMIC)) substantially outperformed GPT-3.5 (AUROC: 0.537, 0.517) and GPT-4 (AUROC: 0.629, 0.602) (with and without in-context learning) in predictive performance and output probability calibration (Brier Score (ML vs GPT-3.5 vs GPT-4): 0.134 versus 0.384 versus 0.251, 0.042 versus 0.06 versus 0.219). Traditional ML is more robust than GPT-3.5 and GPT-4 to generalizing demographic information to protect privacy. GPT-4 is the fairest model according to our selected metrics but at the cost of poor model performance.

**Conclusion:** These findings suggest that LLMs are much less effective and robust than locally-trained ML for clinical prediction tasks, but they are getting better over time.

## INTRODUCTION

Making predictions is a vital part of healthcare, with common applications including estimating the likelihood of a specific diagnosis, assessing the suitability of a medication, and determining a patient’s readiness for discharge. Over the past several decades, the practice of medicine has evolved from clinical intuition to incorporate data-driven guidance, which is increasingly aided by artificial intelligence (AI) and particularly machine learning (ML). While model development has advanced, it is widely recognized that implementing ML models successfully often requires training on or fine-tuning (e.g., via transfer learning (3)) with local, representative data (1,2).

However, not all institutions have sufficient resources to implement ML effectively. For instance, community hospitals may lack the volume of ML-ready data, computational power, and personnel that academic medical centers possess (4). More recently developed ML technologies, such as large language models (LLMs), have hinted at a potential to mitigate these challenges and fundamentally change the integration of ML in medicine. Cloud-based LLMs, such as GPT-3.5 (5), GPT-4 (6) and Claude (7), are pre-trained and can be interacted with conversationally, characteristics that reduce technical friction required to create or use ML (or more broadly, AI) for healthcare settings. While there have been recent investigations into the performance (8–11) and fairness (12) of LLMs versus traditional ML, as well as other diagnostic aids (9,13), a number of critical aspects, including reliability and robustness (e.g., model calibration) of these tools, remain unknown. Given the rapid pace at which LLMs are growing in their production (14), it is critical to quantify their strengths and weaknesses to ensure responsible use.

To learn more about the efficacy of LLMs for clinical prediction, several aspects of real-world implementation need to be considered. First, we need to represent the type of users and constraints that may limit how LLMs are applied in a healthcare setting. In this study, we consider two distinct types of users: 1) the everyday user, whose goal is to retrieve an answer using a single LLM query (i.e., zero-shot prompting), and 2) the advanced user who utilizes retrieval augmented generation (RAG) to contextually present the LLM with similar cases or additional relevant examples in the prompt to augment its decision-making capabilities for better results. In-context learning is such a strategy that helps guide LLM inference (8).

Second, privacy concerns can limit the amount of patient information that can be supplied to an LLM under the control of a third party, such as OpenAI, as these technologies risk exposing protected health information (PHI) to organizations that are neither covered entities nor their business associates as required by the Health Insurance Portability and Accountability Act (HIPAA). Thus, it is likely that patient data would need to be de-identified before submission to the LLM. Moreover, while this may address the privacy requirements of federal regulations, such as the HIPAA Privacy Rule, evidence suggests that as data detail is reduced, the predictive power of the ML is weakened (16). The impact on LLMs, however, remains unclear. Third, many commercial LLMs do not provide detailed documentation of their architecture or training data. As a result, reproducibility and the design of formal evaluation frameworks are limited for prediction tasks in healthcare.

In this study, we evaluate the utility, privacy, and fairness of LLMs compared to traditional ML, using electronic health record (EHR) data from Vanderbilt University Medical Center (VUMC) (17) to predict the likelihood of patient discharge from the hospital within 24 hours and the public use MIMIC-IV and MIMIC-IV ED datasets from Beth Israel Deaconess Medical Center (BIDMC) (18,19) to predict the likelihood of transfer to the ICU within 24 hours after triage in the emergency department. As our investigation shows, traditional ML models significantly outperform LLMs (both with and without in-context learning) in terms of both classification efficacy and calibration. From the privacy perspective, we further find that traditional ML is generally more robust to the reduction in detail of a patient’s demographic information. We also observe that GPT-4 is the fairest model, which may be inappropriately reassuring since GPT-4’s apparent advantage in fairness seems largely a consequence of its poor performance across all subgroups rather than improved performance on disadvantaged subgroups. We did note improvement in performance from GPT3.5 to GPT 4.

## MATERIALS AND METHODS

This study was approved by VUMC’s Institutional Review Board (IRB# 191892).

### Datasets

This study relied on several EHR-derived datasets from VUMC and BIDMC. The MIMIC-IV and MIMIC-IV ED datasets (n = 393,576) contains hospital admission records and emergency department records, respectively, for BIDMC (18–20). From these, we derived a dataset to predict if a patient will be transferred to an intensive care unit (ICU) within 24 hours of hospital admission based on their presentation during Emergency Department (ED) triage. In this dataset, the predictors include race, age, arrival transport type, triage vital signs, and the number of ED, ICU, and inpatient hospital stays for the past 30, 90, and 365 days.

The VUMC dataset (n = 28,880) was developed to predict the likelihood of patient discharge from the hospital within 24 hours(17). We ran preliminary experiments using gradient-boosted trees implemented with CatBoost (21). We predicted likelihood of discharge using the demographic and audit log data features. We then calculated feature importance based on change in the loss function (implemented within the CatBoost library) and selected the 15 most important audit log features. Those features and the demographic features – race, age, current day of week, insurance type, and Area Deprivation Index (22) of residence – were relied upon in this portion of our study.

### GPT

Our experiments were run on a HIPAA-compliant Azure instantiation of GPT to guard PHI. We utilized GPT-3.5 and GPT-4 with temperature set to 0 (to reduce likelihood of extraneous text being generated) and the maximum number of tokens set to 1024. We randomly selected 1,500 data points from each dataset to query LLMs. The features for each dataset were represented in JSON. Supplementary Table S2 specifies specific prompt structures for each dataset. We considered both zero-shot prompting (not providing additional examples) and few-shot examples (see below).

### Retrieval Augmented Generation

To implement RAG, we selected likely relevant examples that would inform LLM inference on the datapoint under consideration. To select instances that are likely to serve as useful in-context examples, we partitioned the dataset *D* into disjoint subsets *D*_1_and *D*_2_. Let *x* be the data point under consideration by the LLM and, without loss of generality, let *x* ∈ *D*_1_. Then, we performed *k*-means clustering on the points in *D*_2_. To select the number of clusters, we measured the inertia (i.e., the sum of squared distances of samples to their closest cluster center (23)) for each value of *k* starting at 2 and continuing until the change in inertia is less than 0.001. We selected the value of *k* based on the Kneedle algorithm, which identifies the point where inertia shows diminishing returns (24). This defined clusters *C*_1_, *C*_2_, …, *C*_*k*_ for *D*_2_. We repeated this process to define clusters for *D*_1_as well such that *k-*means is executed once for *D*_1_ and once for *D*_2_.

With *C*_*i*_ defined as the cluster to which *x* belongs, the data points and their corresponding true labels in this cluster were used as relevant illustrating examples to present to the LLM. Such examples are known as few-shot (or *n-*shot) examples. If *C*_*i*_ had fewer than five members, then the entire cluster was used as few-shot examples; otherwise, we selected five points in *C*_*i*_ with the closest Euclidean distance to *x* (e.g., 5-shot examples). We denote the use of in-context examples as GPT-X RAG Closest Within Cluster (or GPT-X RAG CWC).

### Traditional ML

We used a traditional ML benchmark of a gradient-boosting tree (GBT) implemented with Catboost (21). Categorical data were replaced with an integer corresponding to the index when sorting the possible values alphanumerically. Missing categorical features are replaced with a “?” and included in the sort. Missing numeric features were replaced with −1. We opted for this strategy for missing values since we assume that a clinician interfacing with an LLM may not use statistical imputation procedures. We used default settings for the GBT and performed 10-fold cross-validation to collect the predictions for each data point per test set (i.e., test-set predictions for each data point in dataset).

### Statistical Analysis

Unless otherwise noted, we produced a total of 30 bootstrapped samples of the predictions and report performance metrics as average and 95% confidence intervals. When applicable, we used Welch’s t-test and one-way analysis of variance (ANOVA) test and considered *p <* 0.05 as significant for all statistical tests. For ROC curves and calibration curves, we performed vertical averaging to aggregate results from the bootstrap samples(25). To perform vertical averaging of the graphs, we defined a comprehensive *x*-axis and average the corresponding *y*-values (interpolating *y*-values if necessary). All analysis was performed using Python version 3.11.5, and the ANOVA test was calculated using SciPy library 1.11.4.

### Design of Performance and Calibration Experiments

To measure prediction performance, we used the area under the Receiver Operating Characteristic (ROC) curve (AUROC). We performed vertical averaging (25) across the thirty bootstrap samples to construct the ROC curves. We compared the average AUROC using the bootstrapped samples (not the average AUROC from the vertically averaged curves since vertical averaging of curves interpolation of y-values is required to maintain a consistent x- axis).

Currently, accessing the LLM architecture or training data to assess model reliability or interpretability is not possible. Therefore, evaluating the reliability and applicability of predictions is crucial to ensuring interpretability and trustworthiness. Since, closed-source LLMs such as GPT-3.5 and GPT-4 are black box classifiers, their primary point of interaction is via prompt-based querying and the generated output. In our case, the generated output was the probability of a specific phenomenon (e.g., likelihood of discharge, likelihood of ICU admittance).

In light of this, we used calibration curves (26) and Brier Score (27) to determine how well-calibrated output probabilities are from each model. Calibration curves plot the fraction of positive predictions given a predicted probability. A perfectly calibrated classifier should produce a calibration curve of *y* = *x*. This would indicate that, for all of the predictions assigned a predicted probability *p*, a fraction *p* of these predictions corresponds to the positive class. The Brier Score corresponds to the squared-loss of predicted probabilities to output labels with a range of [0,1], where a lower value indicates a better-calibrated classifier - with 0 indicating perfect calibration and 1 indicating the worst possible calibration. The calibration curves were constructed by vertically averaging (25) calibration curves of the thirty bootstrap samples. Average Brier Score was calculated with the bootstrap samples, using the same technique employed with ROC curves.

We performed Welch’s t-test to determine if there were statistically significant differences in both performance and calibration. Consider *model*_1_ and *model*_2_. The null hypothesis was ℋ_0_: *μ*_*model*_1__ = *μ*_*model*_2__. For AUROC, ℋ_1_: *μ*_*model*_1__ > *μ*_*model*_2__, whereas for Brier Score, ℋ_1_: *μ*_*model*_1__ < *μ*_*model*_2__. Thus, these hypothesis tests determined if *model*_1_ performs better than *model*_2_ for the appropriate metric.

### Design of Privacy-Utility Tradeoff Experiments

To evaluate the privacy-utility tradeoff associated with LLMs, we considered the following patient data obfuscation procedure. Data was organized along three levels of detail as documented in Supplementary Figure S3. Level 0 indicates no data obfuscation (i.e., the original data); Level 1 indicates a mild amount of obfuscation, where each feature was divided into 4-8 categories; and Level 2 indicates moderate obfuscation where each feature was in a binary category (e.g., for race, white/not white). Level 3 indicates all demographic data were suppressed. Real-valued features were partitioned into ranges in Levels 1-3, while categorical features at Levels 1-3 were grouped into semantically similar categories. Sex was suppressed for Levels 1-3. To determine if there was a statistically significant difference in LLM and ML performance between these levels, we performed a one-way analysis of variance (ANOVA) test with a *p*-value threshold of 0.05.

### Design of Algorithmic Fairness Experiments

To assess the fairness of ML and LLMs, we considered subgroups (majority/minority) based on race (white/not white), age (18-49 years old/50 years or older), and sex (male/female). We measured algorithmic fairness in two ways: 1) average absolute odds difference (AAOD) and 2) statistical parity difference (SPD). AAOD is a measure based on the equalized odds (EO) measure of algorithmic fairness (28). EO defines a fair classifier as one for which *TPR*_*s*_*maj*__ = *TPR*_*s*_*min*__ and *FPR*_*s*_*maj*__ = *FPR*_*s*_*min*__ for each possible pair of distinct demographic subgroups *s*_*maj*_ (majority) and *s*_*min*_ (minority). Then, we can calculate AAOD as follows: 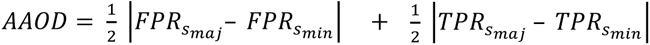. Thus, a perfectly fair classifier will have an AAOD of 0.

SPD measures the difference in the rate that each sensitive group receives a positive prediction, denoted as 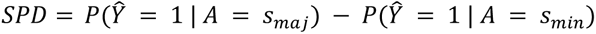 (29). An SPD of 0 corresponds to a perfectly fair classifier. A negative SPD indicates a preference for the minority class, whereas a positive SPD favors the majority class. Since SPD can range from −1 to 1, in addition to calculating the magnitude of statistical parity, we also calculated the absolute value of SPD (|SPD|). For Welch’s T-test, we then have ℋ_0_: *μ*_*model*_1__ = *μ*_*model*_2__ and ℋ_1_: *μ*_*model*_1__ < *μ*_*model*_2__. Since AAOD and |SPD| depict the magnitude of unfairness, smaller metric values indicate higher fairness. We report the sum of the fairness metrics per demographic subgroup.

## RESULTS

### Performance and Calibration

Table 1 summarizes the predictive performance of the LLMs and traditional ML model. As shown in Figure 1, the ROC curves for traditional ML were much larger than those for the LLMs, both with and without in-context learning. Moreover, the p-values in Figure 2 indicated this difference in performance between traditional ML and LLMs was statistically significant. Thus, in terms of measures of discrimination, traditional ML appeared to be more capable at identifying different classes of patients than LLMs at the given clinical tasks. This was not surprising because traditional ML learns a classification function based on a subset of representative data, and it has defined knowledge of expected feature distributions. In-context examples improved the ROC of GPT for all dataset-model combinations except for GPT-3.5 on VUMC data. Nonetheless, traditional ML still outperformed GPT with in-context examples.

**Figure 1.**
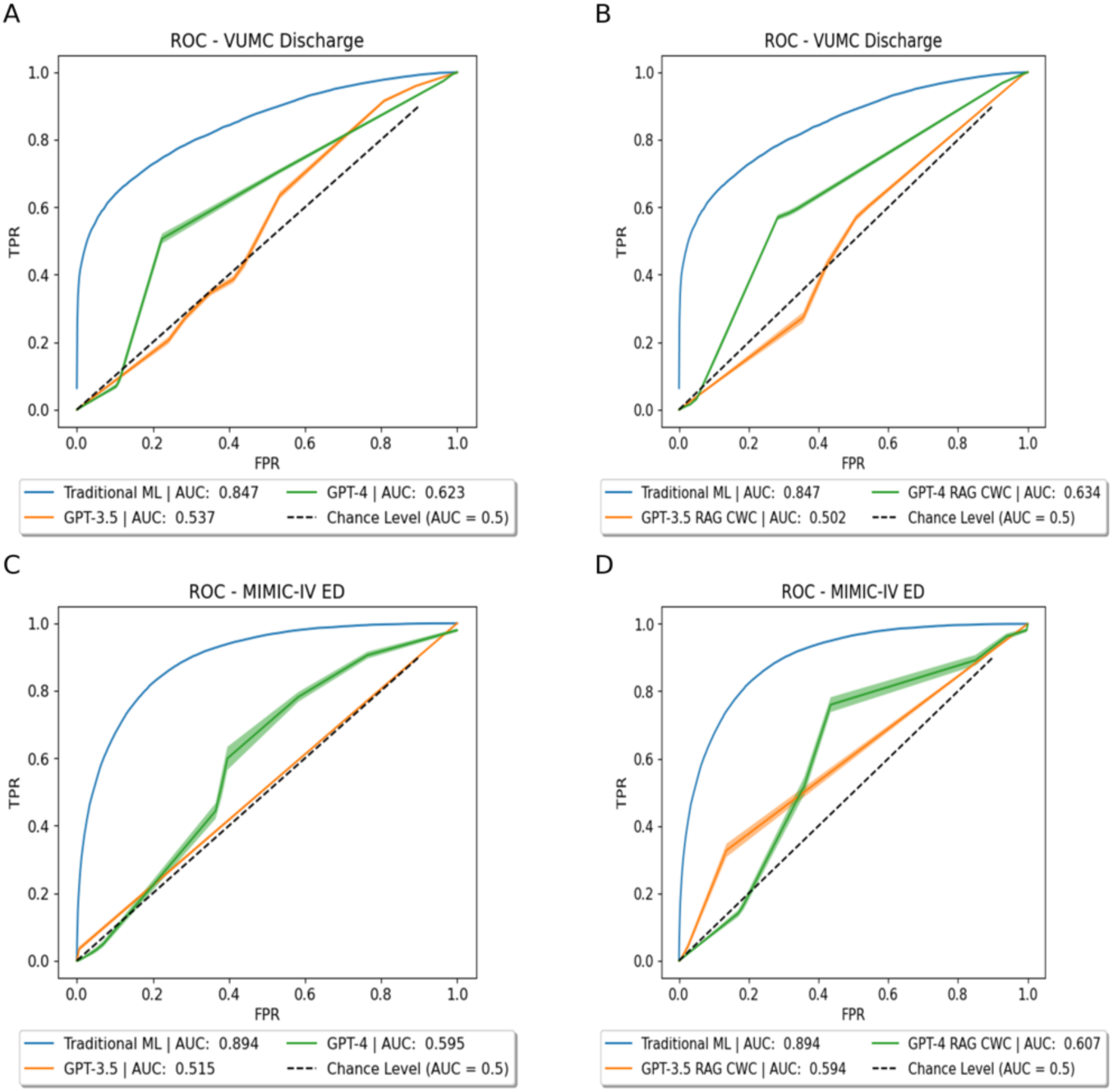
ROC curves for (A) zero-shot prompting on the VUMC dataset, (B) RAG-based few-shot prompting on the VUMC dataset, (C) zero-shot prompting on the MIMIC-IV dataset, and (D) RAG-based few-show prompting on the MIMIC-IV dataset. Note that the AUC given in the legends may not match those given in Table 1 due to numerical interpolation of the curves required for vertical curve averaging (AUC is of the final averaged curve).

**Figure 2.**
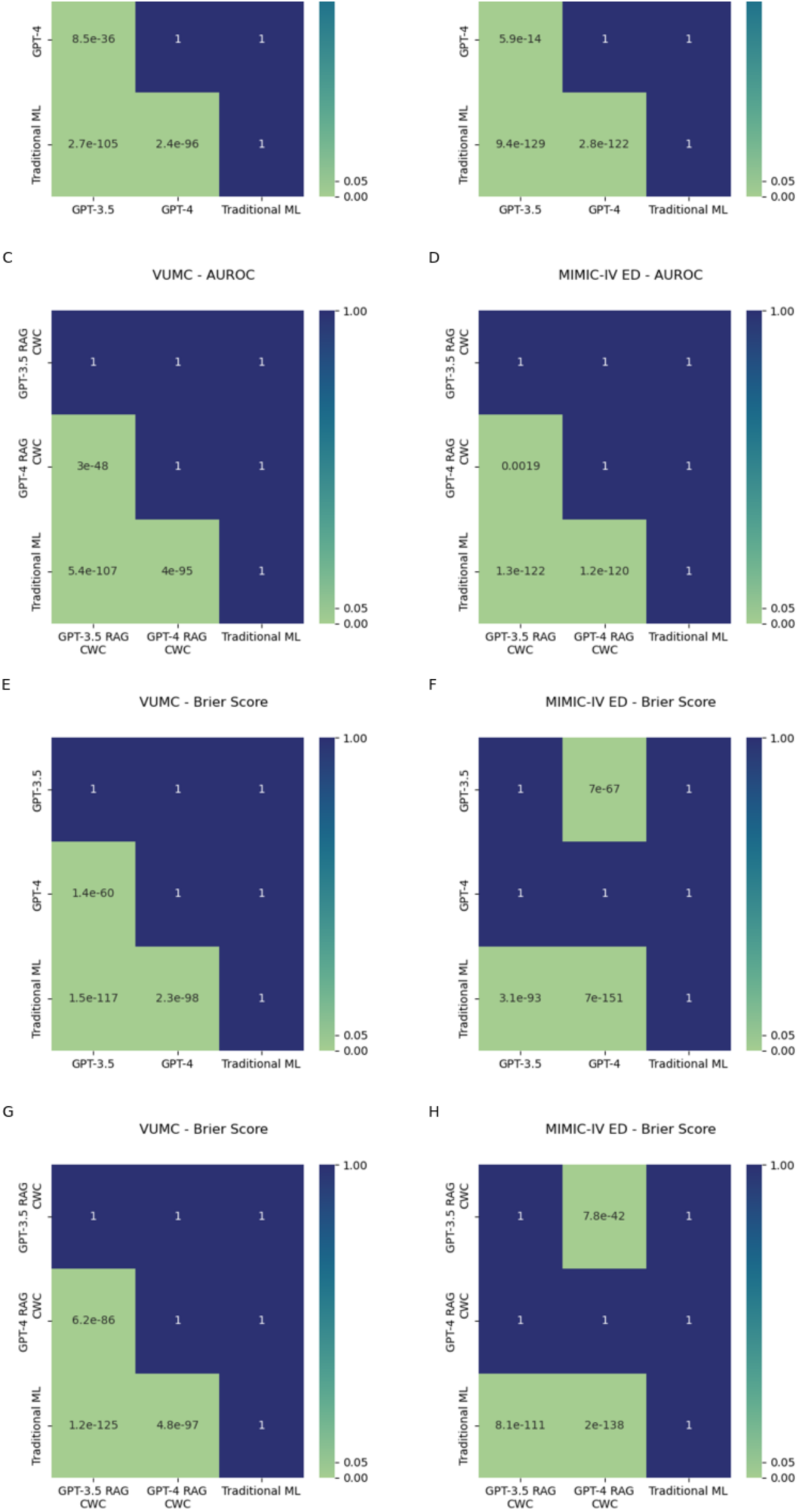
Let *model*_1_ be the model given by the row and *model*_2_ be the model given by the column. Then, ℋ_0_: *μ*_*model*_1__ = *μ*_*model*_2__. For AUROC ℋ_1_: *μ*_*model*_1__ > *μ*_*model*_2__. For Brier Score, ℋ_1_: *μ*_*model*_1__ < *μ*_*model*_2__ Thus, these hypothesis tests determine if *model*_1_ performs *better* than *model*_2_ for the appropriate metric. (A) AUROC from zero-shot prompting on VUMC dataset, (B) AUROC from zero-shot prompting on MIMIC-IV dataset, (C) AUROC from RAG-based few-shot prompting on VUMC dataset, (D) AUROC from RAG-based few-shot prompting on MIMIC-IV dataset, (E) Brier Score from zero-shot prompting on VUMC dataset, (F) Brier Score from zero-shot prompting on MIMIC-IV dataset, (G) Brier Score from RAG-based few-shot prompting on VUMC dataset, (H) Brier Score from RAG-based few-shot prompting on MIMIC-IV dataset

**Table 1.** Average AUROC and Brier Scores and 95% confidence intervals. Arrow indicates better direction. Best in bold.(↑ = higher is better; ↓ = lower is better)

|  | VUMC Discharge |  | MIMIC-IV ED |  |
| --- | --- | --- | --- | --- |
| | AUROC ↑ | Brier Score ↓ | AUROC ↑ | Brier Score ↓ |
| **GPT-3.5** | 0.537 (0.532, 0.543) | 0.384 (0.38, 0.388) | 0.517 (0.513, 0.521) | 0.06 (0.057, 0.062) |
| **GPT-4** | 0.629 (0.625, 0.634) | 0.251 (0.249, 0.253) | 0.602 (0.592, 0.612) | 0.219 (0.216, 0.222) |
| **GPT-3.5 RAG CWC** | 0.512 (0.509, 0.515) | 0.477 (0.47, 0.484) | 0.608 (0.596, 0.619) | 0.078 (0.076, 0.081) |
| **GPT-4 RAG CWC** | 0.638 (0.632, 0.644) | 0.246 (0.244, 0.248) | 0.61 (0.598, 0.621) | 0.152 (0.15, 0.153) |
| **Trad. ML** | **0.847 (0.845, 0.848)** | **0.134 (0.133, 0.134)** | **0.894 (0.894, 0.895)** | **0.042 (0.042, 0.042)** |

Figure 3 depicts the calibration curves for traditional ML, GPT-3.5, and GPT-4. The classification curve for traditional ML was nearly perfectly overlaid the *y* = *x* curve. The calibration curves for GPT-3.5 and GPT-4, however, were heavily miscalibrated. These findings were corroborated by the Brier Score, which can be interpreted as a calibration loss, such that lower Brier scores imply better calibrated classifiers. Traditional ML was better calibrated than GPT-3.5 or GPT-4, a result that was statistically significant, as shown in Figure 2. In-context examples improved calibration for GPT-4 on both datasets, but GPT-3.5 exhibited a decrease in probability calibration for both datasets.

**Figure 3.**
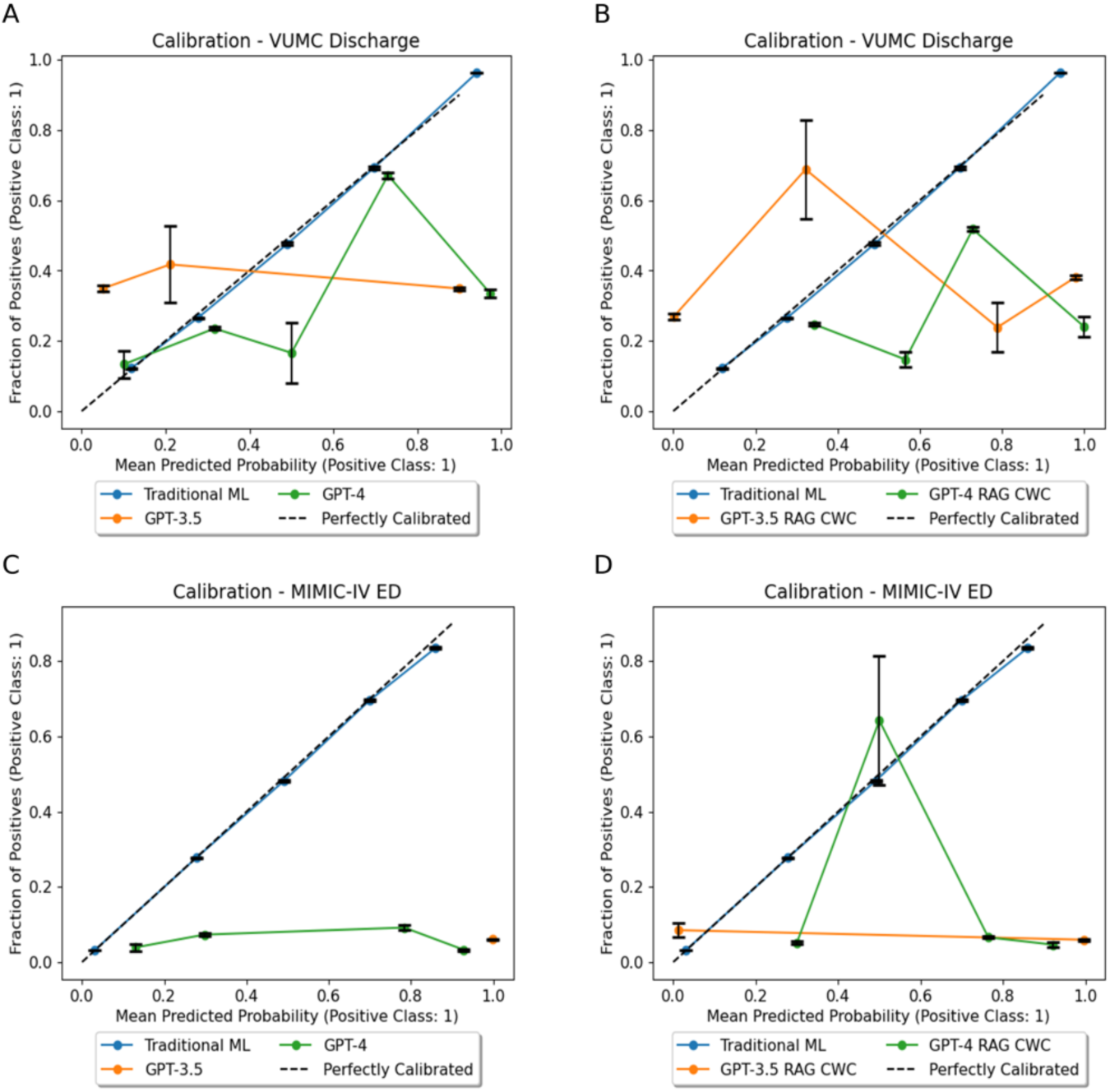
Calibration plots (A) zero-shot prompting on the VUMC dataset, (B) RAG-based few-shot prompting on the VUMC dataset, (C) zero-shot prompting on the MIMIC-IV dataset, and (D) RAG-based few-show prompting on the MIMIC-IV dataset.

### Privacy-Utility Tradeoff

Table 2 presents the mean performance (with 95% confidence intervals in parentheses) of each level of generalization per model. There was less than a 0.01 difference among the standard deviations of performance among different levels of generalization/suppression per model. The ANOVA analysis was statistically significant across all combinations of datasets and models (Table 3), indicating all models were affected, to some degree, by the generalization and suppression of demographic information.

**Table 2.** Average AUROC and 95% CI for each model and level of data obfuscation.

|  |  | VUMC | MIMIC-IV ED |
| --- | --- | --- | --- |
| GPT-3.5 | Level 0 | 0.537 (0.532, 0.543) | 0.517 (0.513, 0.521) |
|  | Level 1 | 0.573 (0.568, 0.578) | 0.526 (0.522, 0.53) |
|  | Level 2 | 0.575 (0.57, 0.579) | 0.515 (0.511, 0.518) |
|  | Level 3 | 0.557 (0.553, 0.561) | 0.514 (0.514, 0.517) |
| GPT-4 | Level 0 | 0.629 (0.625, 0.634) | 0.602 (0.592, 0.612) |
|  | Level 1 | 0.657 (0.652, 0.662) | 0.633 (0.623, 0.644) |
|  | Level 2 | 0.64 (0.636, 0.644) | 0.609 (0.599, 0.619) |
|  | Level 3 | 0.649 (0.644, 0.655) | 0.601 (0.59, 0.611) |
| GPT-3.5 RAG CWC | Level 0 | 0.512 (0.509, 0.515) | 0.608 (0.596, 0.619) |
|  | Level 1 | 0.557 (0.551, 0.562) | 0.631 (0.621, 0.642) |
|  | Level 2 | 0.543 (0.54, 0.546) | 0.605 (0.594, 0.615) |
|  | Level 3 | 0.522 (0.516, 0.527) | 0.564 (0.553, 0.575) |
| GPT-4 RAG CWC | Level 0 | 0.638 (0.632, 0.644) | 0.61 (0.598, 0.621) |
|  | Level 1 | 0.61 (0.605, 0.615) | 0.617 (0.602, 0.631) |
|  | Level 2 | 0.603 (0.598, 0.609) | 0.62 (0.611, 0.629) |
|  | Level 3 | 0.605 (0.599, 0.611) | 0.53 (0.522, 0.538) |
| Traditional ML | Level 0 | 0.847 (0.845, 0.848) | 0.894 (0.894, 0.895) |
|  | Level 1 | 0.844 (0.843, 0.845) | 0.894 (0.894, 0.894) |
|  | Level 2 | 0.845 (0.844, 0.846) | 0.892 (0.891, 0.892) |
|  | Level 3 | 0.846 (0.844, 0.847) | 0.894 (0.894, 0.895) |

**Table 3.** Results of one-way ANOVA analysis to compare the effect of data generalization/suppression on LLM AUROC.

|  | GPT-3.5 |  | GPT-3.5 RAG |  | GPT-4 |  | GPT-4 RAG |  | Traditional ML |  |
| --- | --- | --- | --- | --- | --- | --- | --- | --- | --- | --- |
| Dataset | F | P | F | P | F | P | F | P | F | P |
| VUMC | 48.341 | 2.42E-20 | 101.859 | 2.31E-32 | 45.743 | 1.39E-19 | 40.065 | 7.63E-18 | 17.441 | 2.92E-10 |
| MIMIC | 10.394 | 4.13E-06 | 50.665 | 5.29E-21 | 11.696 | 9.43E-07 | 96.963 | 1.80E-31 | 92.853 | 1.03E-39 |

Figure 4 displays the privacy-utility tradeoff across four levels of demographic generalization and suppression. We observed that traditional ML performance was generally consistent across the levels of demographic generalization and suppression, while GPT-3.5 and GPT-4 displayed varying degrees of performance but no overall directional trend. Next, we considered how LLMs compare to traditional ML as we reduced the granularity (generalize) and remove (suppress) demographic information. By visual inspection, traditional ML was relatively more robust to generalization and suppression than LLMs. LLMs varied more in performance, with little noticeable trend in movement. Still, when demographic information was removed, LLMs (and particularly LLMs with in-context learning) dropped in performance. This raises the concern that LLMs may have some level of reliance on demographic information when performing zero-shot predictions.

**Figure 4.**
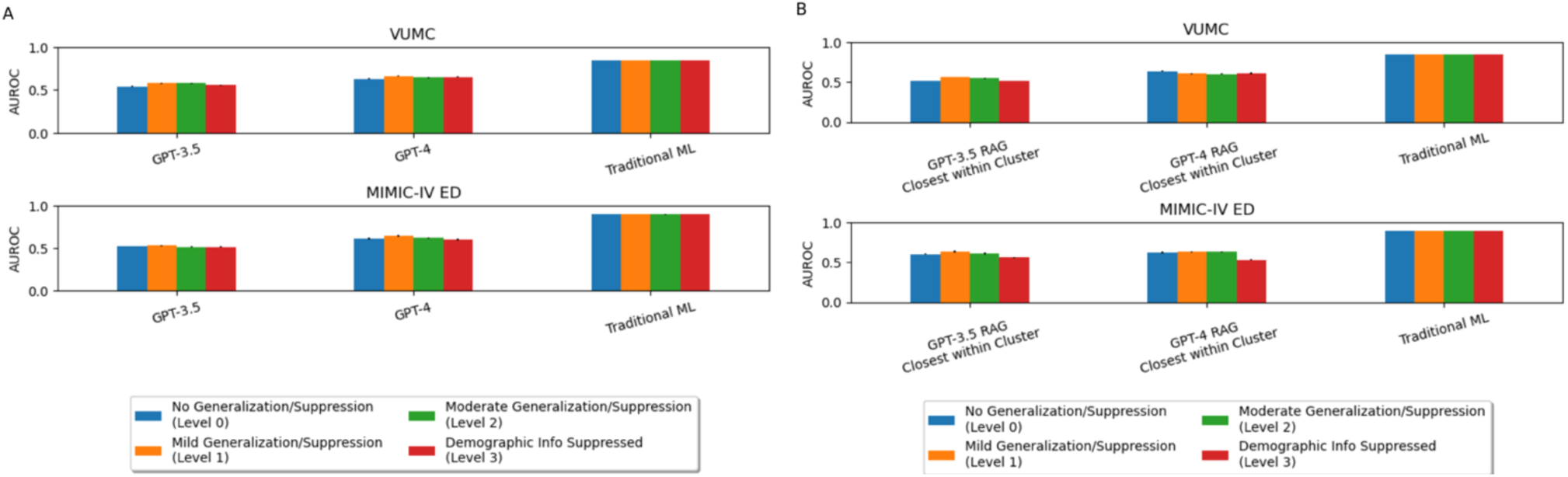
Bar charts depicting the privacy-utility tradeoff for (A) zero-shot prompting and (B) RAG-based few-shot prompting.

### Algorithmic Fairness

Figures 5 and 6 plot model overall performance and fairness on an XY-plane. It was observed that traditional ML was not unambiguously the fairest model. For the VUMC data, GPT-4 with in-context examples was the fairest model across both fairness metrics, while in-context examples did not improve GPT-3.5 fairness. For the MIMIC dataset, GPT-4 was the fairest model, but in-context examples improved the fairness of GPT-3.5.

**Figure 5.**
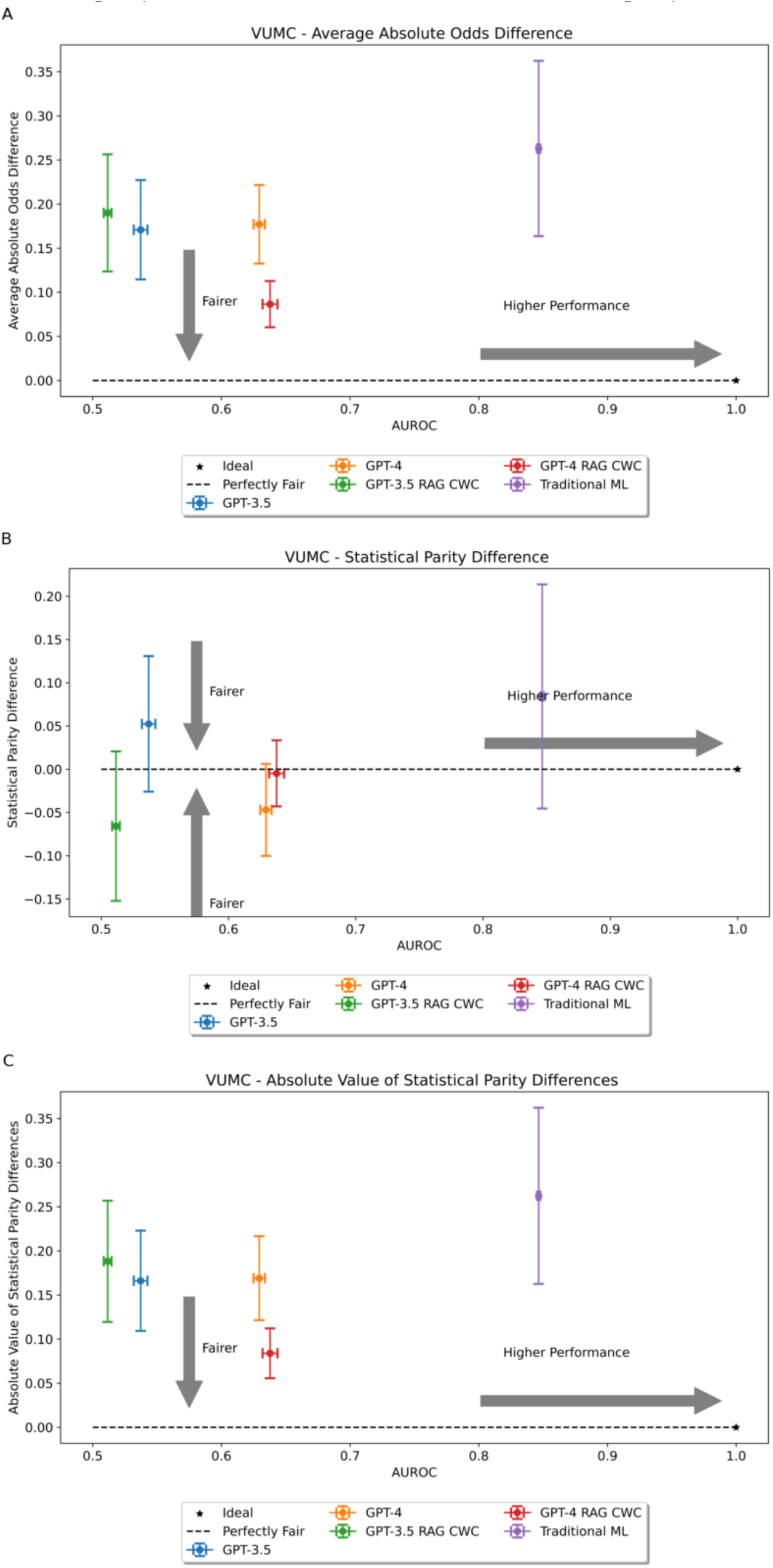
Two-dimensional fairness plots for the VUMC dataset using (A) average absolute odds difference, (B) statistical parity difference, or (C) absolute value of statistical parity difference as the fairness metric, respectively.

**Figure 6.**
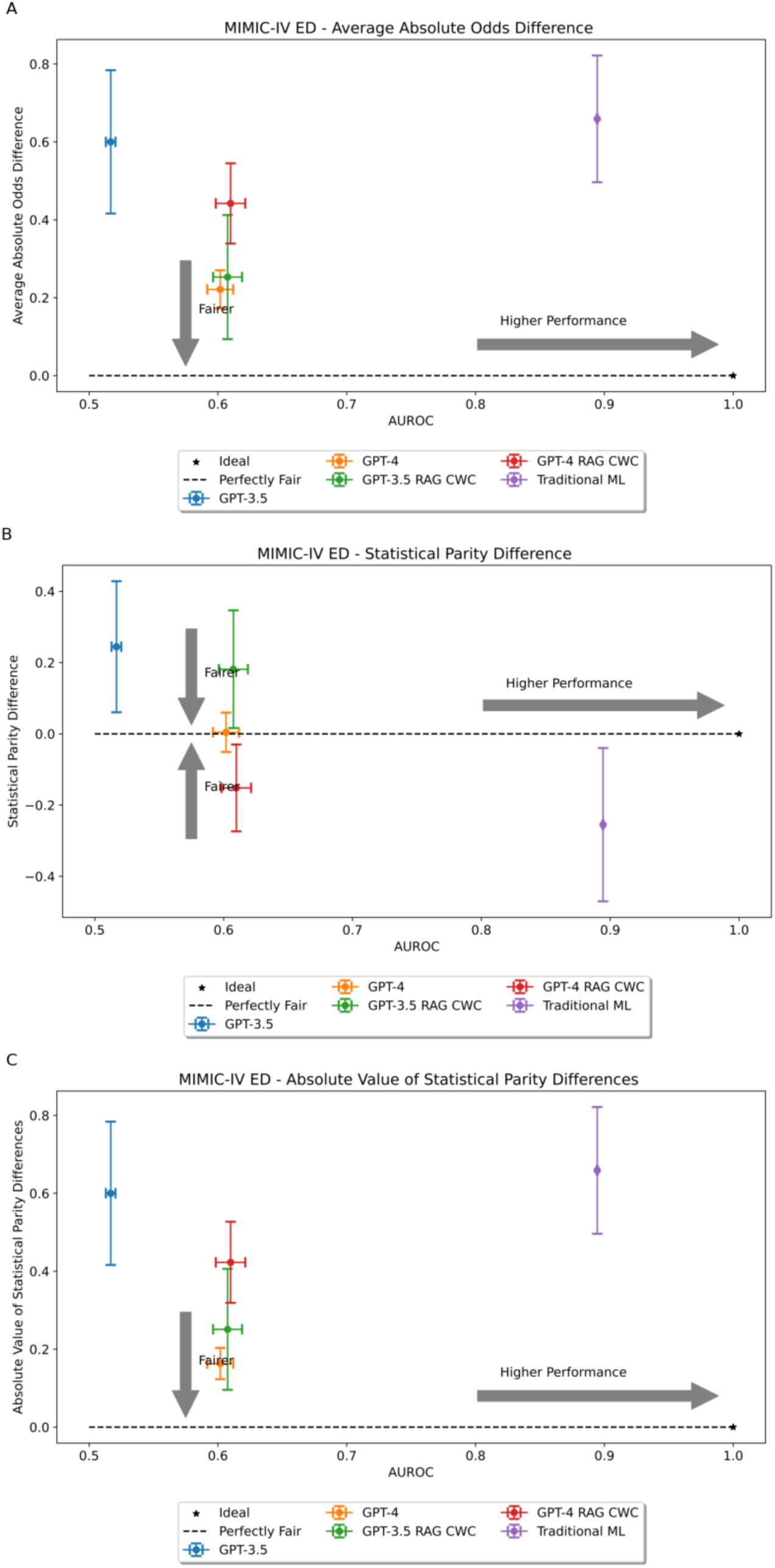
Two-dimensional fairness plots for the MIMIC-IV dataset using (A) average absolute odds difference, (B) statistical parity difference, or (C) absolute value of statistical parity difference as the fairness metric, respectively

We further evaluated the statistical significance of differences in fairness metrics between different models by subgroup. Supplementary Figure S4 provides heatmaps of p-values from the corresponding Welch’s t-tests determining statistical significance of the results. There are twelve possible scenarios where one model could be statistically significantly fairer than another (based on demographic variable, fairness metric, and dataset). For all twelve cases, GPT-4 was statistically significantly fairer than traditional ML; however, incorporating in-context examples reduced this number to ten of twelve cases. For GPT-3.5, zero-shot learning was statistically significantly fairer than traditional ML for only six of twelve cases. In the VUMC dataset, GPT-3.5 struggled with maintaining fairness for race, but in the MIMIC dataset, GPT-3.5 was only fairer for subgroups defined by sex. In-context learning did not help the fairness for the VUMC data; however, in-context examples improved GPT-3.5’s fairness for the race and age variables in the mimic dataset. When comparing GPT-3.5 and GPT-4, we note that there is no unambiguously fairer model. Without in-context examples, GPT-4 surpassed GPT-3.5’s fairness for all demographics in the MIMIC dataset but none in the VUMC dataset. With in-context examples, GPT-4 was fairer on the VUMC dataset, yet GPT-3.5 still surpassed GPT-4’s fairness on the MIMIC dataset. We note that this fine-grained analysis may differ somewhat from the overall trends presented in Figures 5 and 6, since these figures consider aggregate fairness across all demographic subgroups considered. As a result, the small, yet significant shifts may be cancelled out in the aggregate.

While GPT-4 was the fairest model for both datasets, the fairer model between GPT-3.5 and traditional ML depended upon dataset and fairness metric considered. Traditional ML was trained and evaluated on data from the same location. Thus, whatever biases are present in the data source will be present in the resulting model. We evaluated GPT-3.5 and GPT-4 using zero-shot prompting and without knowledge of the evaluation datasets. Thus, these models were not exposed to the local dataset biases that may influence decisions. This led us to hypothesize that traditional ML would be the most unfair model, which was validated by our findings. The use of in-context examples also impacted fairness; however, this impact was not unanimously positive or negative but depended upon dataset and metric. Moreover, despite being the fairer models, GPT-3.5 and GPT-4 were also the least capable predictors. Thus, it seems that overall improved fairness comes at the cost of reduced subgroup performance; but future work is necessary to confirm.

## DISCUSSION

This investigation suggests that LLMs are not yet ready to serve as predictive, analytic models, although GPT-4 does surpass GPT-3.5 in most of our experiments, indicating that GPT is improving with new releases. In our experiments, LLMs have a significantly lower AUROC compared to traditional ML. This implies that LLMs simply do not match the discriminative capabilities of traditional ML for these tasks. Moreover, inspecting the calibration curves and Brier Scores reveals that LLMs are poorly calibrated when compared to the traditional ML comparison. The weak calibration of the LLM predictions in clinical scenarios calls into question their utility and reliability as a clinical classifier. This is not to say LLMs do not have advantages as a classifier, as there is little model tuning required out-of-the-box to be minimally functional (whereas traditional ML requires training data and computation time) and the conversational interaction may allow for chain-of-thought reasoning to support usability and interpretability (30). These advantages, however, are difficult to realize when the output probabilities do not indicate reliability. Unlike several traditional ML algorithms (31,32), LLMs have limited capability to quantify their uncertainty post-hoc (33). Thus, if LLMs are going to be useful as accurate clinical predictors, further research into improving the reliability and interpretability of LLMs is a must.

For LLMs to be useful as classifiers in lower resource settings, for example, where extensive data may not be available to train models, strategies such as in-context learning and RAG are necessary to produce good performance. We expected the incorporation of intelligently selected in-context examples (8,34) would improve LLM performance to levels near that of traditional ML. Instead, we find that in-context examples do not always improve LLM discriminative performance. Table 1 reveals that for the VUMC data, the usage of in-context examples resulted in either a slight increase in performance (GPT-4) or a decrease in performance (GPT-3.5). Further research is needed to improve selection of RAG-based in-context examples for biomedical classification.

In the event that LLMs are the most accessible or applicable model for a situation, there are still concerns with respect to patient privacy when using these models. As we alluded to, organizations may apply some form of de-identification before sending data to the LLM, but our analysis indicates that LLMs are much more sensitive to the level of detail in demographic information compared to traditional ML. This suggests that a strategy that generalizes patient information may result in a reduction in predictive performance.

The apparent sensitivity of LLMs to the completeness of patient demographic information raises questions about the biases and unfairness present in these models based on their existing training. We find that GPT-4 is the fairest model evaluated; however, we note that LLMs are generally the lowest performing models evaluated. By definition, an unfair model is one that results in a disparate outcome for at least one subgroup (35). Typically, mitigating fairness concerns usually means improving the performance of the disadvantaged subgroup(s). This route ensures equity of performance with reduced harm (36). For the LLMs, increased fairness seems to come at reduced performance for all subgroups. This is evidenced by the overall degradation of performance of LLMs compared to traditional ML. Thus, it may be difficult to justify the use of LLMs on the basis of apparent fairness in the context of subpar performance.

Moreover, we find that there is not a consistently fair model. Fairness is conditioned on dataset and prediction task. For example, work from Liu et al. (12) also compared the fairness of GPT-3.5 to traditional ML. In their analysis, traditional ML was fairer in their analysis when compared to GPT-3.5 in two of the three evaluated datasets. In our evaluation, traditional ML is fairer than GPT-3.5 for age and gender subgroups in the MIMIC dataset; however, this does not hold for the VUMC dataset or race in the MIMIC dataset. Liu et al. also found that fine-tuning using few-shot examples improves the fairness gap for GPT-3.5, but there is little discussion on the performance impact. Our analysis indicates that few-shot examples can improve fairness of GPT-3.5 and GPT-4, but whether or not this occurs is dependent on model and dataset.

Though we conducted a large number of experiments, we acknowledge several limitations of this investigation. First, other facets of evaluation beyond the scope of this work, such as interpretability, were not evaluated and could also differ between model types. Second, we did not consider the costs of implementing and maintaining these models and the associated environmental factors. Operational costs of AI may prohibit lower-resourced medical centers from utilizing these advances, and the energy required may have a downstream negative impact both economically and environmentally. Third, we only consider two clinical tasks from datasets derived from two medical centers. Additional studies that consider a wider variety of tasks across multiple medical centers is crucial for generalizability of these findings. Finally, we note that the privacy-utility tradeoff reported does not match the trade-off expected. Ultimately, this calls into question the reliability of the datasets used for this privacy-utility tradeoff analysis. We cannot rule out the possibility that information captured in other predictors in these datasets are serving as appropriate proxies to demographic information.

## CONCLUSION

This study reported on a multifaceted comparison of LLMs to traditional ML in terms of model performance, calibration, privacy, and fairness as the points of evaluation. Even when supplemented with in-context learning, we find that LLMs are not as effective as traditional ML for clinical prediction tasks due to their poor discriminative performance and lack of reliability, particularly when demographic factors are variables in the prediction model. While LLMs are the fairest technologies, they are also those with poor model performance. This indicates that performance across subgroups degrades to ensure fairness but this needs to be confirmed.

Still, we note that LLMs have potential. Since March 2023, LLMs have been used to aid with drafting patient messages (37), optimizing clinical decision support (38), developing phenotyping algorithms (39), and summarizing patient encounters (40). In the next eighteen to twenty-four months, LLMs may be better optimized to serve as clinical prediction models. To reach this goal, research is needed to optimize in-context learning for LLM performance, understand the effect of randomization-based privacy preservation strategies, and understand the internal biases of LLMs to mitigate fairness concerns.

## Data Availability

The datasets generated and/or analyzed during the current study are not publicly available due to patient private information investigated but are available from the corresponding authors on reasonable request.

## ACKNOWLEDGEMENTS

This research was sponsored, in part, from NIH grants U54HG012510 and T15LM007450, from NSF grants IIS-1905558

## AUTHOR CONTRIBUTIONS

Using the CRediT Taxonomy, the following are the roles of the authors on the manuscript:

KB: Conceptualization, Formal analysis, Investigation, Writing (original draft, review & editing)

CY: Conceptualization, Writing (original draft, review & editing)

ZL: Writing (review & editing)

XZ: Data curation, Writing (review & editing)

BC: Writing (review & editing)

YC: Data curation, Writing (review & editing)

EC: Writing (review & editing)

MK: Conceptualization, Writing (review & editing)

YV: Conceptualization, Writing (review & editing)

BM: Conceptualization, Resources, Supervision, Writing (original draft, review & editing)

