## Supplemental Files for "Not the Models You Are Looking For: Traditional ML Outperforms LLMs in Clinical Prediction Tasks"

### S.1 Additional Dataset Information

This section details information about the evaluated datasets from BIDMC and VUMC. This includes the feature name, feature type, possible values (if categorical), and the unit and normal range provided to GPT.

Table 1. Detail for the features in each dataset. Italicized features are considered demographic variables.

| **Dataset** | **Feature Name** | **Type w/o generalization/suppression** | **Values** | **Unit & Normal Range provided to GPT** |
|---|---|---|---|---|
| MIMIC | No. Days in the ED in the past <30/90/365> days | Integer | | |
| | No. Days as an Inpatient in the Hospital in the past <30/90/365> days | Integer | | |
| | No. Days in the ICU in the past <30/90/365> days | Integer | | |
| | *Age* | Float | | Years |
| | *Gender* | Categorical | Male/Female | |
| | *Race* | Categorical | White, Asian, Black/African-American, Hispanic/Latino, Native Hawaiian or Other Pacific Islander, Multiple Race/Ethnicity, Patient Declined to Answer, ? | |
| | Temperature at Triage | Continuous | | degrees Fahrenheit (Normal: 97-99 degrees Fahrenheit) |
| | Heartrate at Triage | Continuous | | beats per minute (Normal: 60-100 beats per minute) |
| | Respiratory Rate at Triage | Continuous | | breaths per minute (Normal: 12-20 breaths per minute) |
| | Oxygen Saturation at Triage | Continuous | | % (Normal: 97%-100%) |
| | Systolic Blood Pressure at Triage | Continuous | | mm Hg (Normal: 90-120 mm Hg) |
| | Diastolic Blood Pressure at Triage | Continuous | | mm Hg (Normal: 60-80 mm Hg) |
| | Reported Pain Level at Triage | Continuous | | Reported Pain Level at Triage (Possible Values Range: 0 = no |

| | | | | |
|---|---|---|---|---|
| | | | | pain, 10+ = severe pain) |
| | Emergency Severity Index | Categorical | 1 (most severe), 2, 3, 4, 5 (least severe) | (Possible Values Range: 1 = most severe, 5 = least severe) |
| VUMC | *Race* | Categorical | Non-Hispanic White, Non-Hispanic Black, Hispanic, Other Race/Ethnicity, Multi | |
| | *Age* | Continuous | Age of patient | Years |
| | *Sex* | Categorical | Male/Female | |
| | *Day of Week* | Categorical | Monday, Tuesday, Wednesday, Thursday, Friday, Saturday, Sunday | |
| | *Insurance* | Categorical | Type of insurance held (Private, self-pay, public) | |
| | *Area Deprivation Index of Residence* | Continuous | ADI of ZIP code listed as residence | |
| | Viewed Discharge Instructions | Continuous | Audit Log Information | Accesses |
| | Order Reconciliation Section accessed | Continuous | | Accesses |
| | Obstetrics redirected print group viewed | Continuous | | Accesses |
| | Patient device removed | Continuous | | Accesses |
| | OR report printed GetSurgicalRecord webservice accessed | Continuous | | Accesses |
| | Report with patient data viewed | Continuous | | Accesses |
| | Communication sent | Continuous | | Accesses |
| | Report with patient data printed | Continuous | | Accesses |
| | Notes viewed | Continuous | | Accesses |
| | Order sets accessed | Continuous | | Accesses |
| | After Visit Summary Viewed | Continuous | | Accesses |
| | Printing occurred | Continuous | | Accesses |
| | Patient based RW report exported | Continuous | | Accesses |

### S.2 LLM Prompt Information

This section details the specific prompt template used for both tasks. We provide the system message to help contextualize the LLM to the task and the question posed to the LLM. Input features are provided in JSON after the system message and before the question.

Table 2. LLM Prompt Template.

| **Dataset** | **Prompt Component** | **Prompt Text** |
|---|---|---|
| VUMC | System Message | You are an expert at predicting patient discharge from a hospital given their demographic information and audit log statistics for the past 24 hours. |
| | Question | Question: Will the patient be discharged on the described day?<br>Provide your answer in the following format: {"answer": 0(no)/1(yes), "predicted_probability": (probability that answer is correct, in decimal, round to three decimal places)}<br>DO NOT produce any additional text. |
| MIMIC | System Message | You are an experienced attending physician who gives precise forecasting of emergency department patient outcomes based on patient presentation at triage. |
| | Question | Question: Will the patient be admitted to an ICU within 24 hours?<br>Provide your answer in the following format: {"answer": 0(no)/1(yes), "predicted_probability": (probability that answer is correct, in decimal, round to three decimal places)}<br>DO NOT produce any additional text. |

### S.3 Data Generalization Diagrams

This section details the hierarchical generalization diagrams for both datasets. Unless otherwise noted, the bottom row depicts the feature value without generalization or suppression. Ascending levels depict a higher level of generalization until the features are suppressed, denoted by *.

Figure S1. Hierarchical generalization diagram for VUMC dataset.

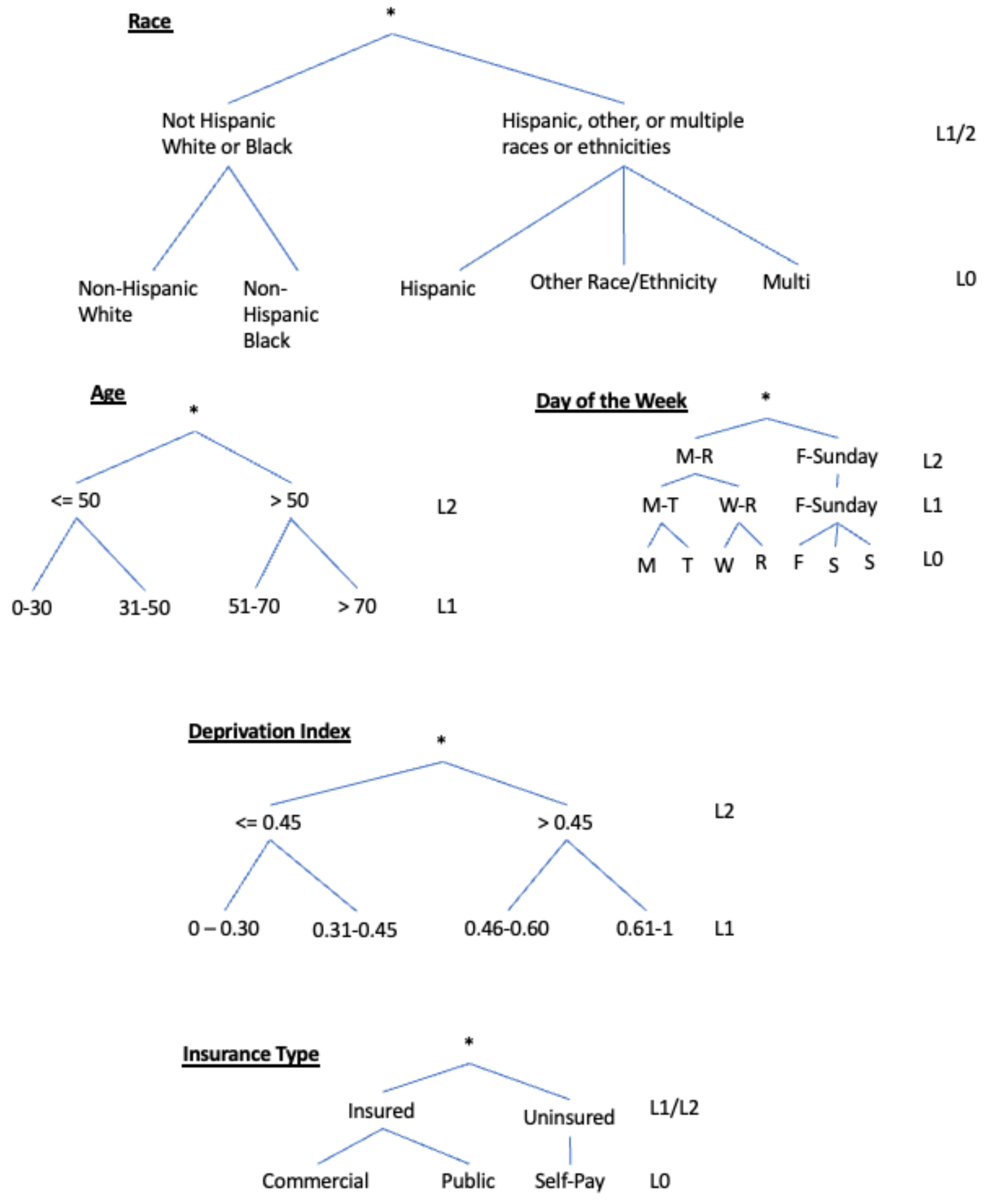

Figure S2. Hierarchical generalization diagram for MIMIC-IV dataset.

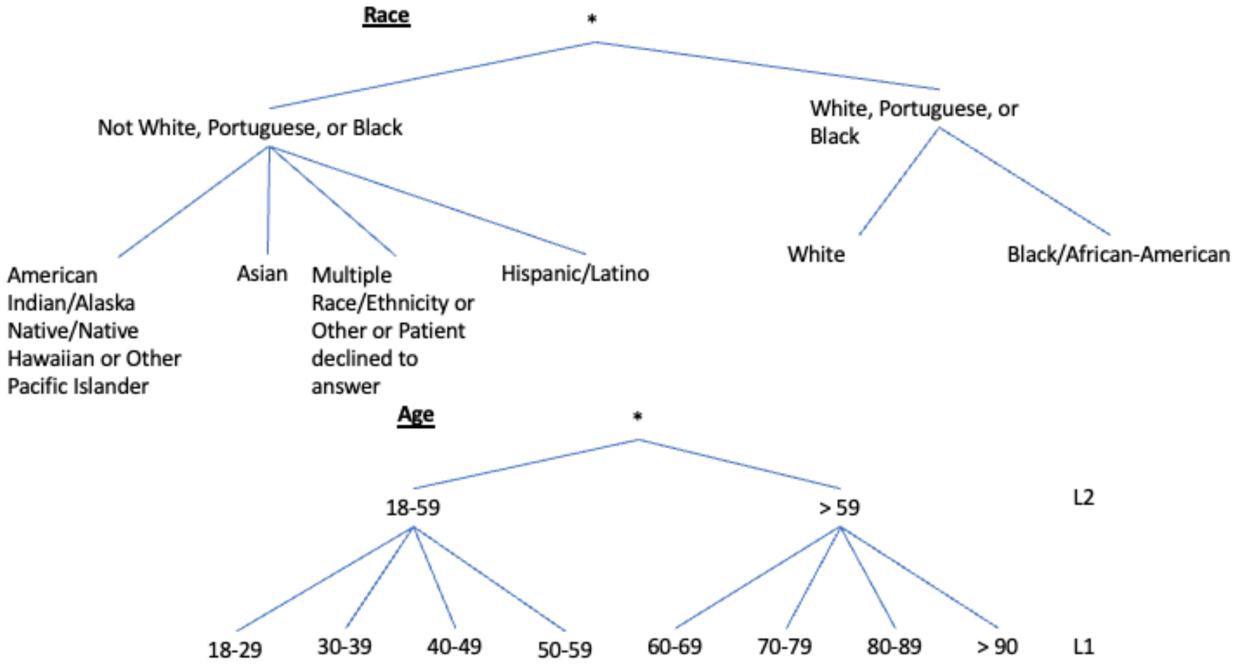

### S.4 Additional Results

This section details additional results from the fairness analysis. This includes heatmaps of the p-values from Welch's t-test to evaluate which model is fairer under average absolute odds difference or absolute value of statistical parity difference. We consider $p < 0.05$ as statistically significant.

Figure S3. This figure is for the VUMC data using (A) average absolute odds difference on zero-shot prompting, (B) average absolute odds difference on RAG-based few-shot prompting, (C) absolute value of statistical parity difference on zero-shot prompting, (B) absolute value of statistical parity difference on RAG-based few-shot prompting. Let $model_1$ be the model given by the row and $model_2$ be the model given by the column. Then, $\mathcal{H}_0: \mu_{model_1} = \mu_{model_2}$ and $\mathcal{H}_1: \mu_{model_1} < \mu_{model_2}$. A lower p-value indicates $model_1$ is fairer (AAOD, |SPD|) than $model_2$.

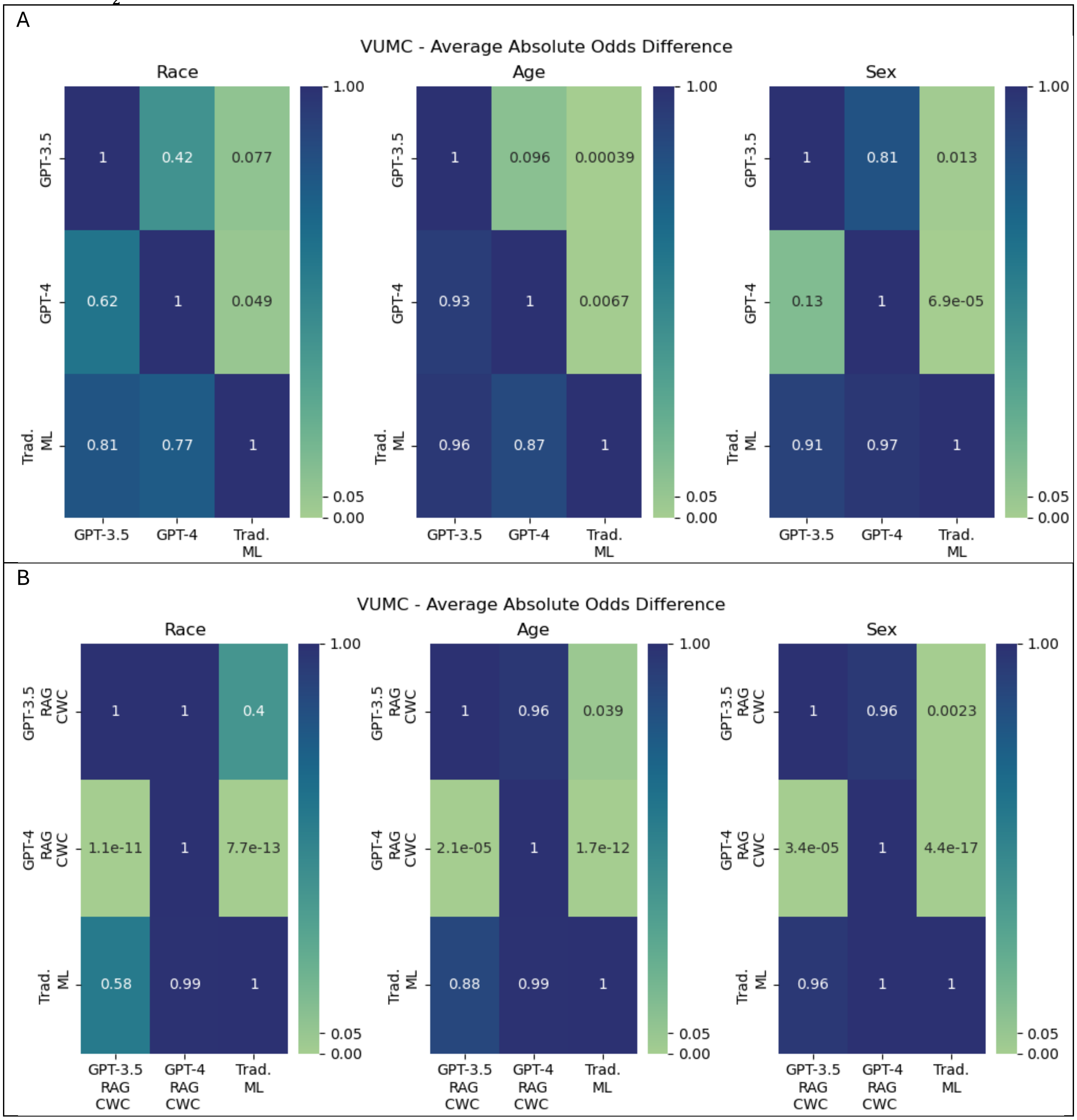

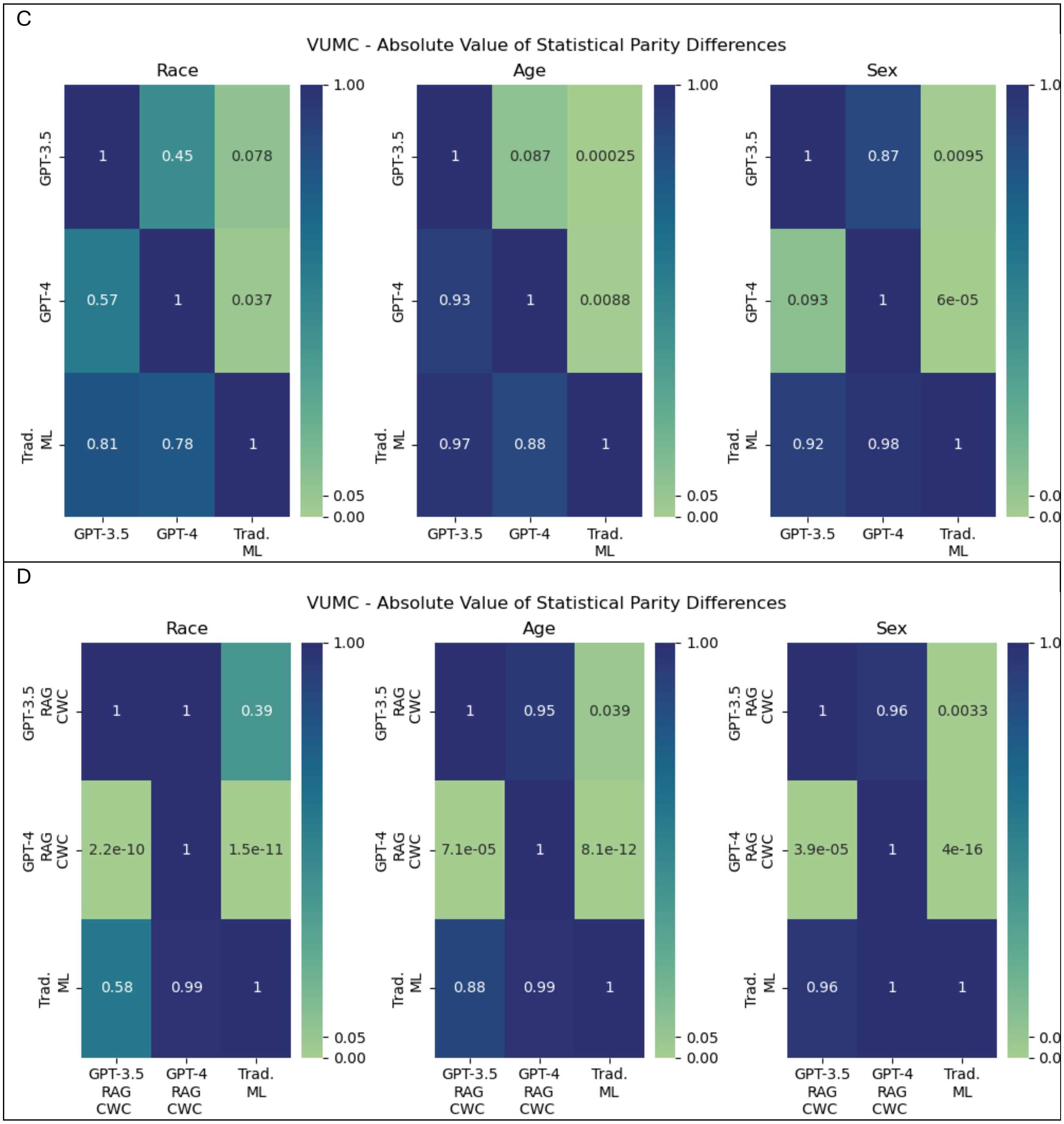
C
VUMC - Absolute Value of Statistical Parity Differences
Race
Age
Sex
GPT-3.5
GPT-4
Trad. ML
1
0.45
0.078
0.57
1
0.037
0.81
0.78
1
0.087
0.00025
0.93
0.0088
0.97
0.88
0.87
0.0095
0.093
6e-05
0.92
0.98
1.00
0.05
0.00
D
VUMC - Absolute Value of Statistical Parity Differences
Race
Age
Sex
GPT-3.5 RAG CWC
GPT-4 RAG CWC
Trad. ML
1
1
0.39
2.2e-10
1
1.5e-11
0.58
0.99
1
0.95
0.039
7.1e-05
8.1e-12
0.88
0.99
0.96
0.0033
3.9e-05
4e-16
0.96
1.00
0.05
0.00

Figure 4. This figure is for the MIMIC-IV data using (A) average absolute odds difference on zero-shot prompting, (B) average absolute odds difference on RAG-based few-shot prompting, (C) absolute value of statistical parity difference on zero-shot prompting, (B) absolute value of statistical parity difference on RAG-based few-shot prompting. Let $model_1$ be the model given by the row and $model_2$ be the model given by the column. Then, $\mathcal{H}_0: \mu_{model_1} = \mu_{model_2}$ and $\mathcal{H}_1: \mu_{model_1} < \mu_{model_2}$. A lower p-value indicates $model_1$ is fairer (AAOD, |SPD|) than $model_2$.

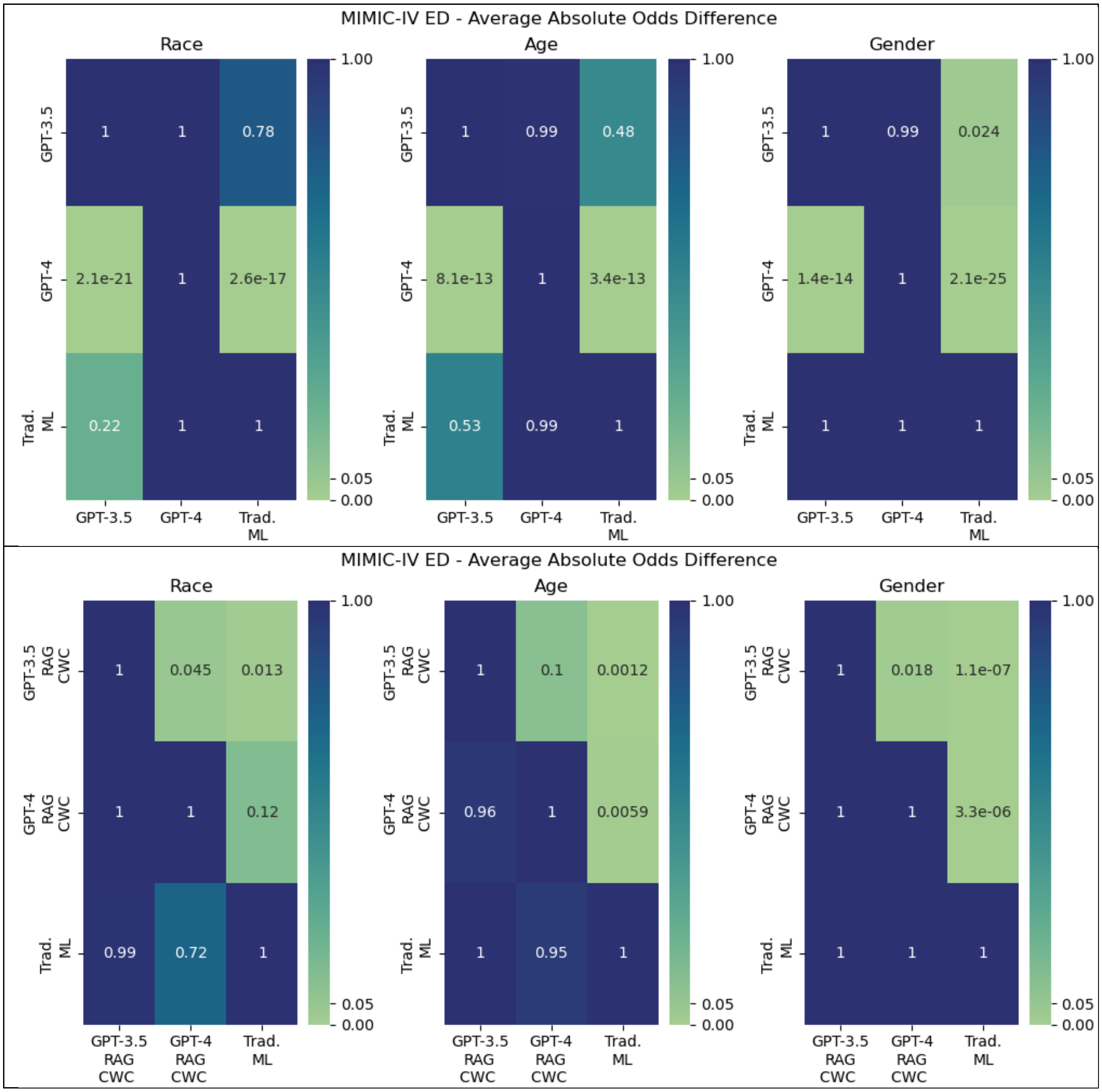

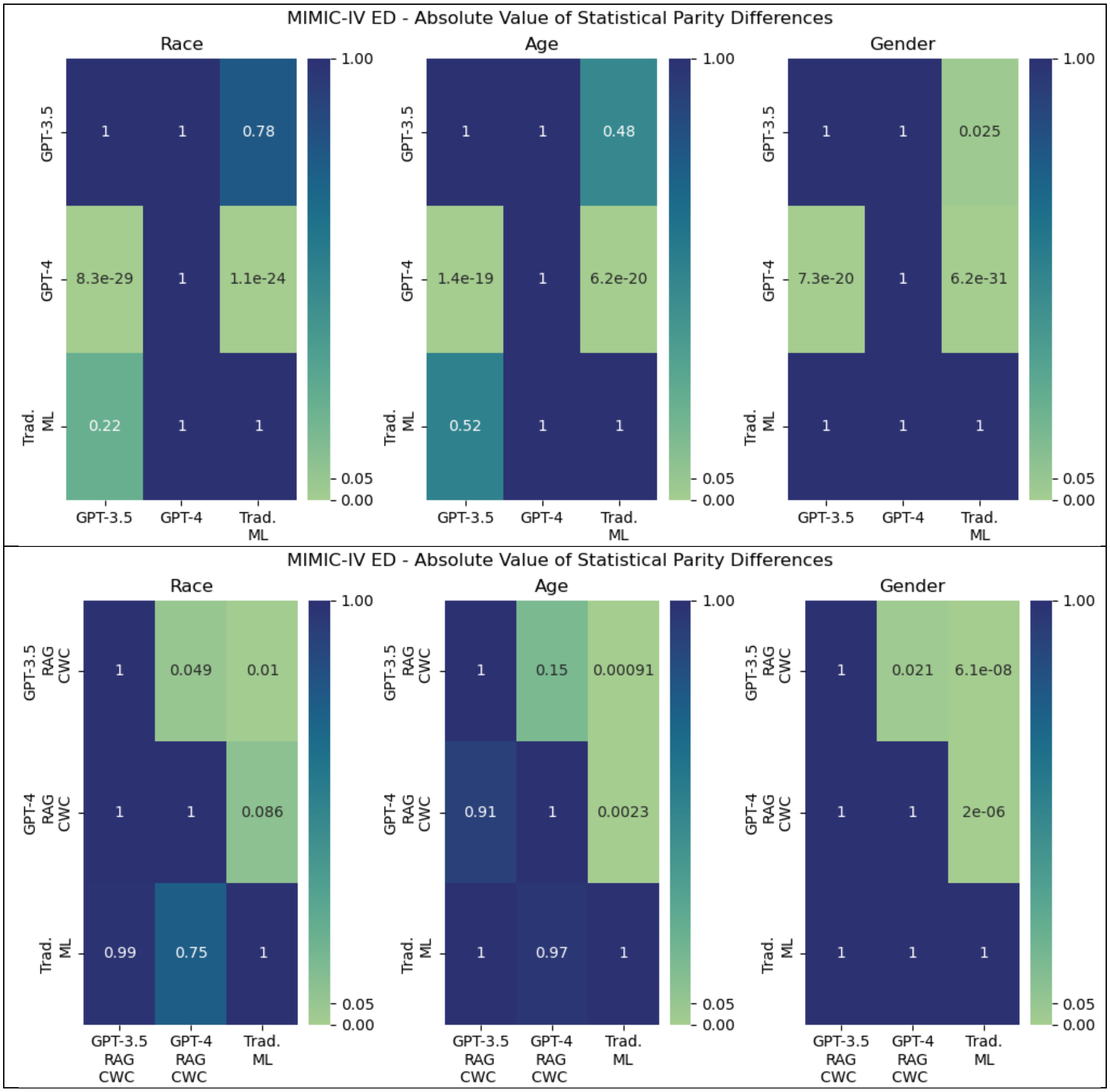
MIMIC-IV ED - Absolute Value of Statistical Parity Differences
Race
Age
Gender
GPT-3.5
GPT-4
Trad. ML
1
0.78
8.3e-29
1.1e-24
0.22
0.48
1.4e-19
6.2e-20
0.52
0.025
7.3e-20
6.2e-31
1.00
0.05
0.00
MIMIC-IV ED - Absolute Value of Statistical Parity Differences
Race
Age
Gender
GPT-3.5 RAG CWC
GPT-4 RAG CWC
Trad. ML
1
0.049
0.01
0.086
0.99
0.75
0.15
0.00091
0.91
0.0023
0.97
0.021
6.1e-08
2e-06
1.00
0.05
0.00
